# Is there evidence that BCG vaccination has non-specific protective effects for COVID 19 infections or is it an illusion created by lack of testing?

**DOI:** 10.1101/2020.04.18.20071142

**Authors:** Shivendu Shivendu, Saurav Chakraborty, Agnieszka Onuchowska, Ankit Patidar, Arpit Srivastava

**Affiliations:** University of South Florida; University of Louisville

## Abstract

The goal of this paper is to showcase that the COVID-19 disease pattern is evolving and to study the relationship between mandatory BCG policy and caseload/million or death/per million. We analyze seven recent publications on the impact of BCG vaccinations on the development of COVID19 illness and extend presented findings using the latest data from April 10, 2020. We analyze data from 98 countries and we extend existing models by adding the dimension of COVID-19-related testing conducted by the analyzed countries. Similarly to prior studies, we find that COVID-19 attributable case and death incidences across countries share a relationship with a country’s BCG vaccination inclusion in the national immunization program when testing is not taken into consideration. However, this relationship vanishes when we add the dimension of testing. We observe that case and death incidences conditional on testing do not get affected by the countries’ BCG vaccination inclusion in the national immunization program. Therefore, we show that there is no statistical evidence to support the assertion that inclusion of BCG vaccination in national immunization program (NIP) has any impact of COVID 19 infections (cases) or mortality.

## Introduction

COVID-19 pandemic has impacted more than 200 countries and territories with at least more than 2.35 million infections and 160,000 fatalities since the beginning of 2020.^1^ Though the disease has attained global spread, its impact in terms of number of infections and deaths per million population across countries has been significantly heterogeneous. While the disease was pre dominantly impacting Asian region around beginning of March 2020, in next six week or so, it has moved to Europe and North American region (Figures 1 and 2). The growth rates of infections and deaths per million population has also been significantly heterogeneous across continents (Figures 3 and 4).

**Figure 1:**
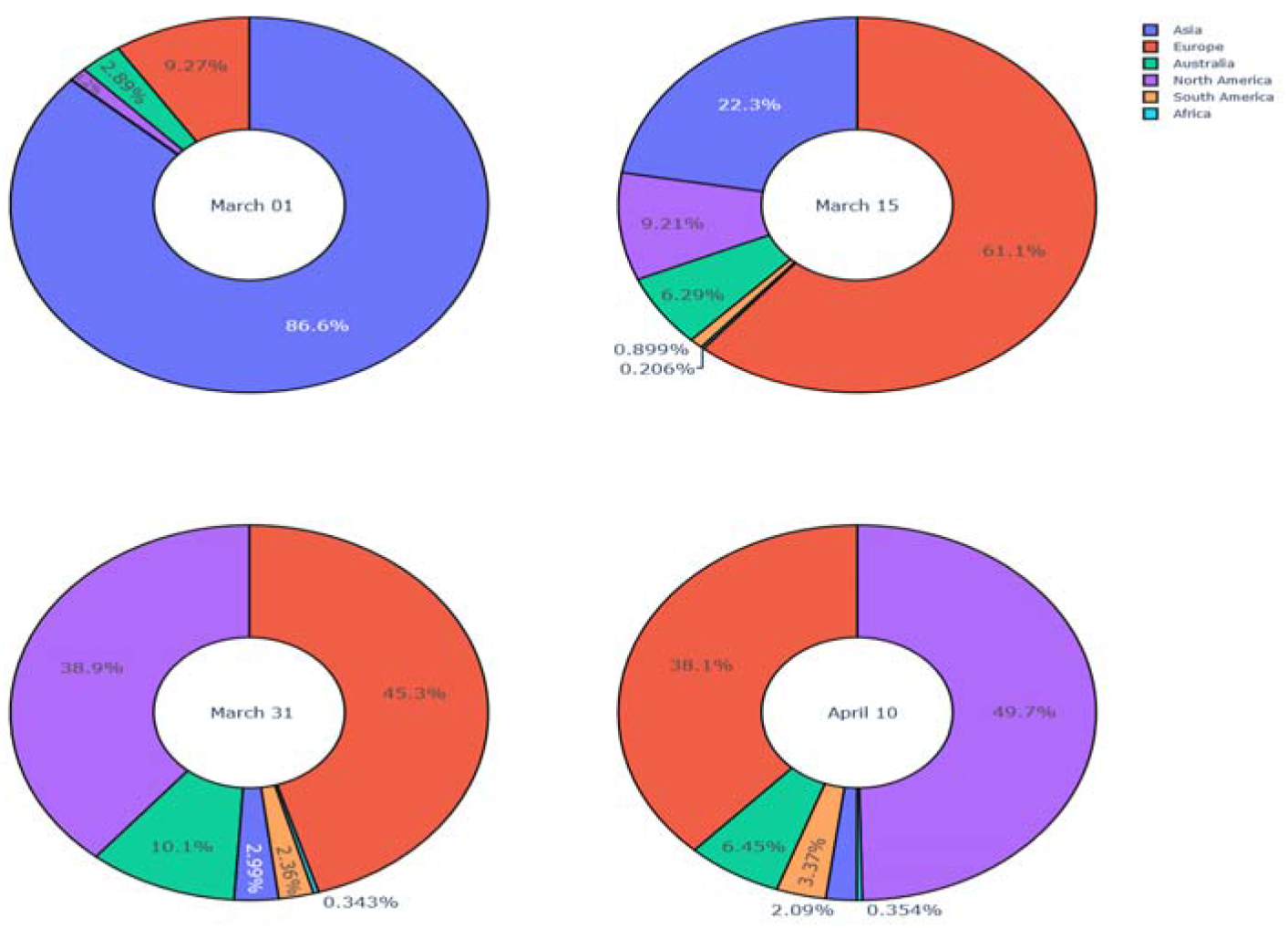
**Case incidence per million population across continents since beginning of March 2020**

**Figure 2:**
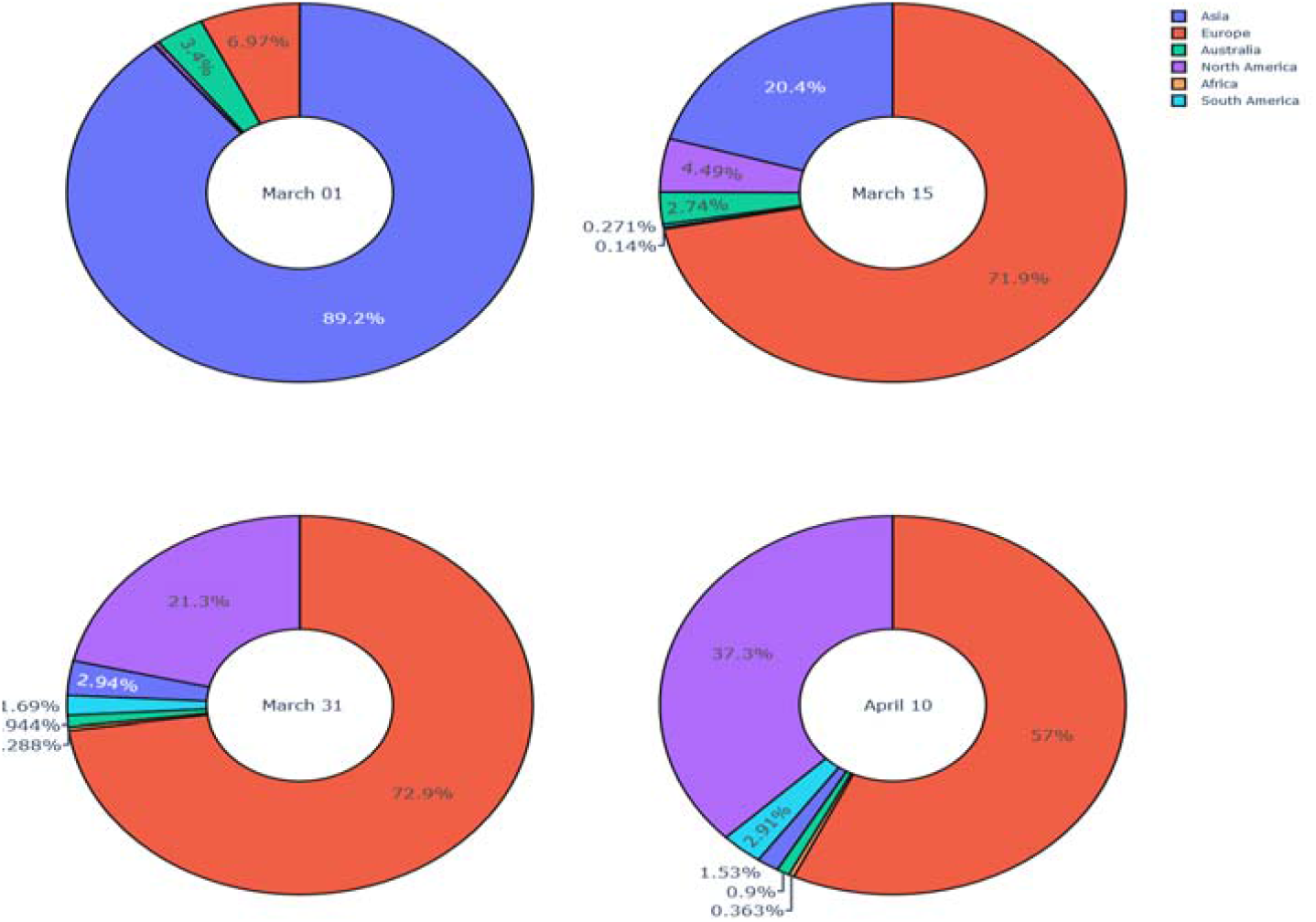
**Mortality per million population across continents since beginning of March 2020**

**Figure 3:**
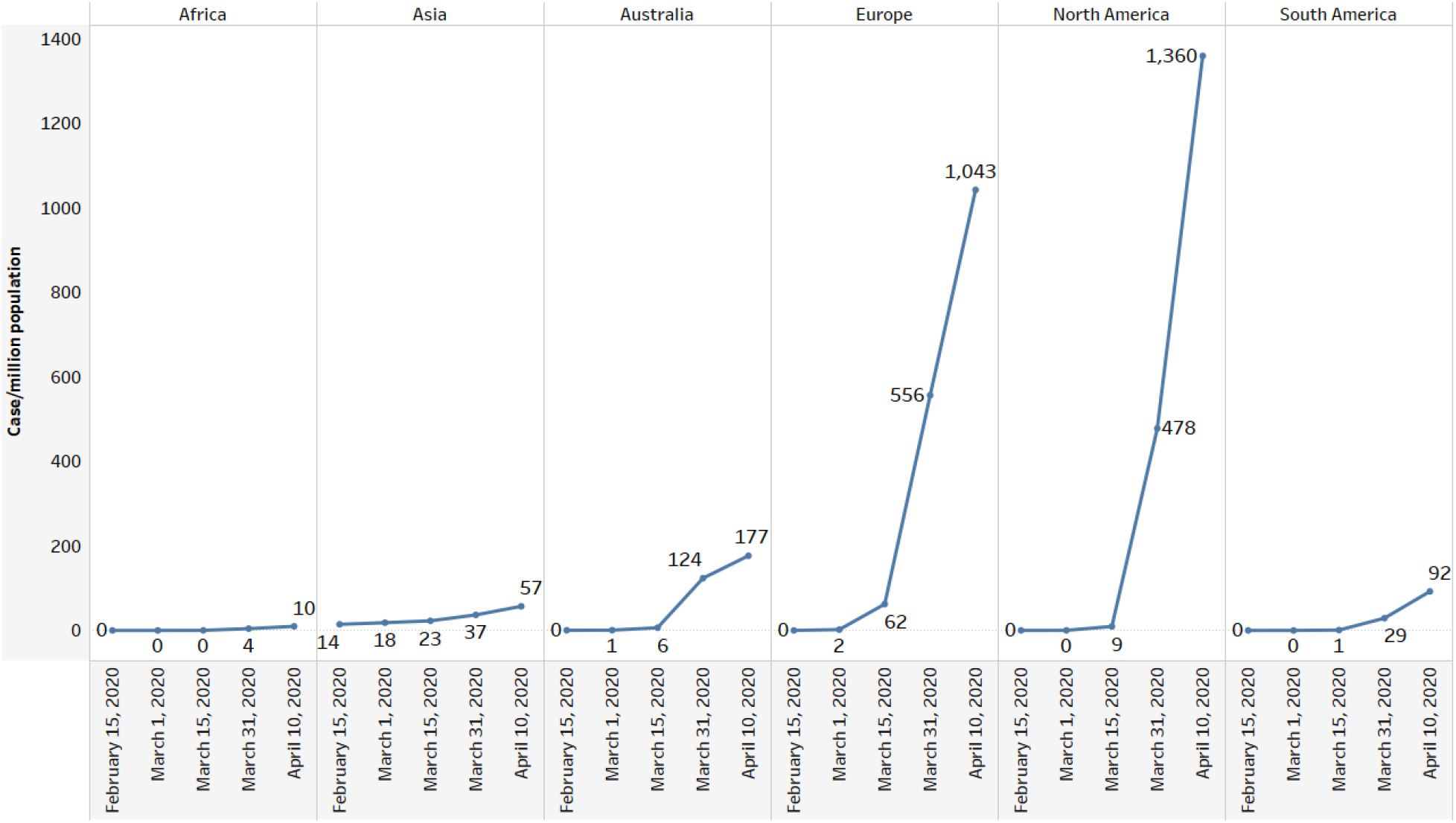
**Evolution of case incidence per million population across continents since February 15, 2020**

**Figure 4:**
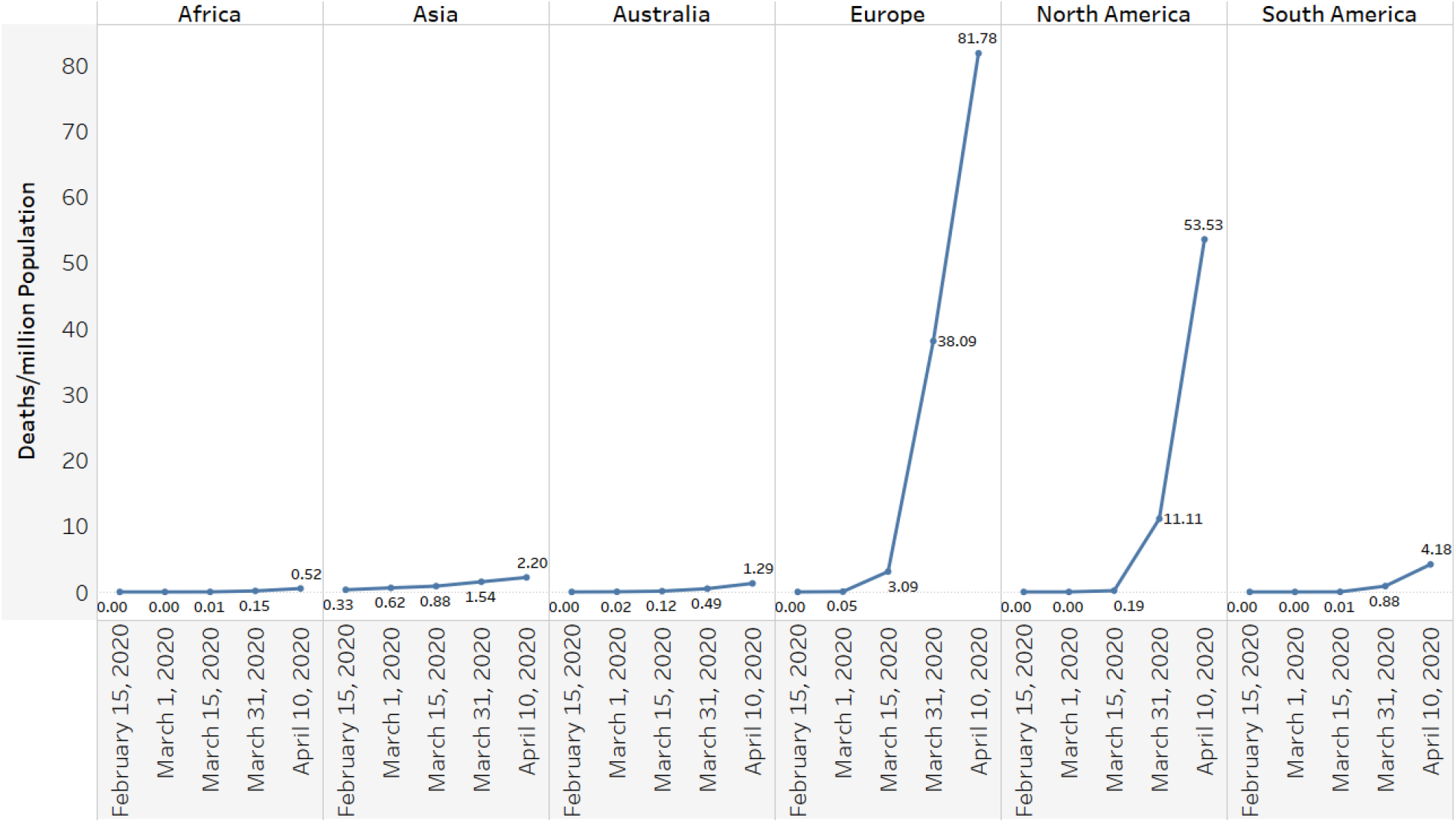
**Evolution of mortality per million population across continents since February 15, 2020**

While there have been significant efforts to understand the science and epidemiology of COVID-19 (Novel 2020; Fauci et al 2020; Wang et al 2020) and also to use various disease modeling approaches to forecast the disease burden as well as healthcare infrastructure requirements in coming months (Hellewell et al 2020; Grasselli 2020; COVID et al 2020; Kucharski et al 2020), there have also been attempts to understand as well as explain noticeable differences across countries/regions in disease incidence, spread and mortality through exogenous factors such as climatic zones (Sajadi et al 2020; O’Reilly et al 2020) and prevailing health policies across countries (Gates 2020; Anderson et al 2020). Within this emerging stream of literature that examines the impact of exogenous factors on COVID-19 spread and outcomes, there has been growing interest in examining the impact of Bacille Calmette Guérin (BCG) vaccine on country level outcomes (Berg et al. 2020; Shet et al 2020; Hegarty et al 2020; Miller et al 2020). This has been of particular interest to health policy community because there has been some evidence to show that BCG vaccine has non-specific protective effects on infections, as well as long term efficacy against tuberculosis (Fine 1995; Dockrell & Smith 2017), and therefore, there may also be some positive non-specific immunity related effects for COVID 19 in BCG vaccinated populations.

Given that there is overwhelming evidence to suggest that COVID-19 is spreading rapidly and its country level patterns are highly dynamic, it is useful in itself to estimate the impact of BCG vaccination on COVID 19 case incidence and mortality with more current data. To our knowledge the most recent study related to our work has used April 01 data (Berg et al 2020). Moreover, none of the previous studies have either controlled for COVID-19 test intensity or have looked at the impact of test intensity on observed dependent variable of cases per million or deaths per million. This is critical, because the dependent variable of interest, either cases per million or deaths per million are observed or measured conditional on potentially infected individual which is detected only through testing of symptomatic or even non-symptomatic individual.

For example, suppose countries A and B have each 1 million population and, at a particular time, have same 10% population infection rate and 10% mortality rate for those who get infected. If country A conducts 100,000 tests, then detected number of infections or cases will be 10,000 and number of deaths attributed to infection would be 1,000. On the other hand, if number of tests in country B are only 10,000, then in country B number of cases detected will be 1,000 and number of deaths attributed to infection would be 100. It is easy to see that while both countries have same infection and mortality rates, country A will report 10,000 cases and 1000 deaths per million, while country B will report only 1,000 cases per million and 100 deaths per million. Therefore, considering cases per million or deaths per million as a measure of infection rate and its severity across countries assumes that the disease detection rates across countries are comparable. On the other hand, data from various countries suggests otherwise: tests per million population vary from 13 (Afghanistan) to 100,002 (Iceland) across countries as of April 10, 2020^2^. Furthermore, data suggest that those countries which are reporting higher number of cases as well as deaths per million are also often times reporting higher number of tests per million (Figure 5).

**Figure 1:**
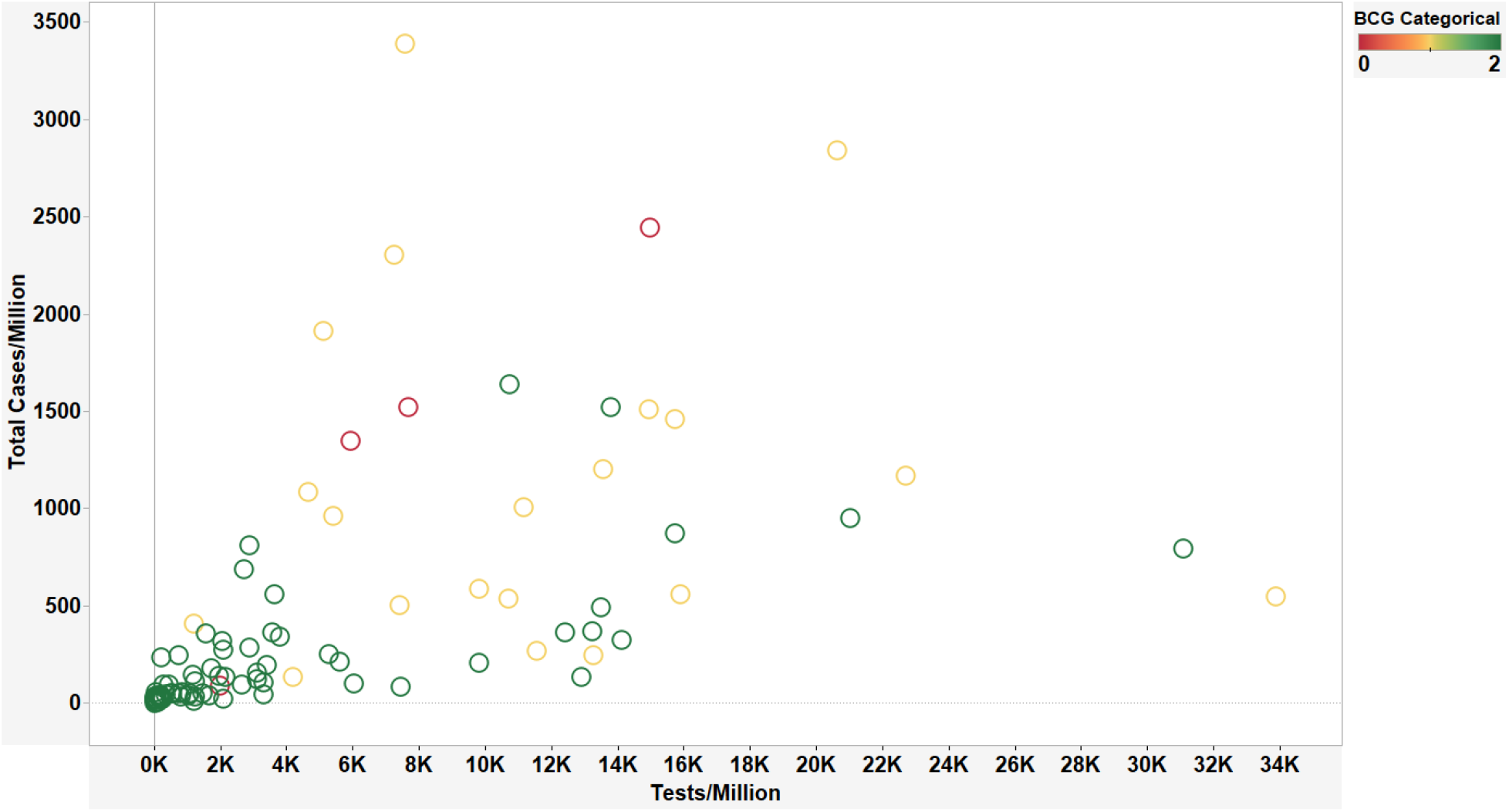
**Tests per million against Cases per million population for countries as of April 10^th^**

The research goal of this paper is to extend the research that empirically seeks evidence to study relationship between BCG vaccination policies of a country (and in turn BCG vaccinated proportion of population) and COVID 19 burden (both in terms of cases and fatalities) at country level. Our work is different from previous studies along four dimensions: (a) while previous studies have used BCG as a categorical variable, we create a variable, BCG Index, which approximates the proportion of population of a country vaccinated against BCG; (b) we introduce additional controls such as HDI score^3^, number of days since one case per million, population density per km^2^, percentage of population above 65 years of age, percentage of population living in urban areas and government transparency score via Corruption Perception Index (CPI)^4^ ; (c) we take into consideration number of tests per million and also use cases per test as dependent variable in the analysis; and (d) use the recent data to verify if the relationships found in earlier studies with prior period data still holds.

To evaluate the effect of BCG vaccine inclusion in country’s national immunization program (NIP) on the COVID-19 infection and mortality levels, we created multiple regression models.

We adjusted for different economic and demographic factors such as GDP per capita, HDI score, percentage of urban population, percentage of population above 65 years of age and other factors such as days since a country reported 100 confirmed COVID-19 cases. We created BCG inclusion variable as a three-tiered categorical variable: countries which have never included BCG vaccine in their NIP, countries which had included the vaccine in the past but do not do so currently, and countries who currently have BCG vaccine included in their NIP. In addition we also created a continuous variable, BCG Index, which captures the number of years a country has included BCG vaccine in its NIP. This index, in our view, is more robust in capturing the coverage of BCG vaccine within a country’s population as compared to a one time snapshot of BCG vaccine inclusion in the NIP for the country.

We evaluated the effect of BCG vaccine on different aspects of infection and mortality within a country. We were especially interested in analyzing how the effect changes when we take testing into account. We observed that there is a considerably high level of heterogeneity across countries when we take into account the total number of tests conducted by them per million population. Furthermore, in order to take into account the incidence of COVID 19 based on testing, we created a dependent variable, case per test, which is created by dividing the total number of cases per million in a country by the number of tests conducted by a country per million population.

Consistent with prior studies, we find that inclusion of BCG vaccine in a country’s NIP has a significant impact in lowering the number of cases per million and deaths per million for that country. Though we observe that the estimated coefficients are smaller in value from previous studies (Shet et al 2020), when we use more current data. However, after taking into account testing intensity, we find that the inclusion of BCG vaccine in NIP has no significant impact on either COVID 19 cases per million or death per million. The results are consistent for both categorical BCG vaccine inclusion variable and the BCG Index. We argue that here may be endogenous factors such as testing that affect the relationship shared between the inclusion of BCG vaccine in a country’s NIP and the number of COVID-19 attributable cases or mortality within the country.

Prior studies may not have taken testing data into consideration due to non-availability of country level data at one source. Worldometer.info/coronavirus/ has been publishing this data since April 4. This provides an opportunity for researchers to consider this critical information in their analysis.

The landscape of COVID-19 attributable cases and deaths throughout the world is constantly evolving. Given the rapidly evolving nature of this disease, we have to be careful before making assertions on how BCG vaccinations influence the emergence of COVID-19-related cases and deaths. Empirical evidence of any beneficial non-specific immunity against COVID 19 infections due to BCG coverage may give a false sense of innate security and protection against the disease to policy makers, health professionals and citizens. Since all most all the developing countries or least developed contries have mandatory BCG coverage in their NIPs and these are also the very countries that have weak public health infrastructure, any false sense of security has the potential to lure them in lowering their efforts to safeguard against COVID 19, including ramping up testing.

We posit that it may be beneficial to create a longitudinal study that can be used to get more insights of the relationship between the BCG vaccine and COVID-19. We are currently in the process of creating a panel dataset to analyze the effects of the BCG vaccine inclusion on COVID-19 attributable cases and mortality rates change over time while controlling for testing intensity. We also propose to release new version of the paper every week based on updated data.

### Literature Review

In order to find research articles that investigate the relation between COVID-19 and BCG vaccinations, we first run a search using Google Scholar search engine. We used keywords ‘BCG and COVID-19’ and restricted the search year to 2020. We found seven non peer-reviewed articles published in late March and early April 2020. Five papers were uploaded onto MedRxiv platform (Berg et al., 2020, Dayal and Gupta, 2020, Miller et al., 2020, Sala and Miyakawa, 2020 and Shet et al., 2020), one study was posted on ResearchGate (Hegarty et al., 2020) and one study became available directly on SSRN (Li et al., 2020). We present a summary the papers in Table 1 and discuss them below.

**Table 1:** Selected literature on COVID-19 and BCG policy relation studies.

| Author | Dataset Characteristics and Timespan | Methodology | Variables of interest | Control variables | Findings/ Results |
| --- | --- | --- | --- | --- | --- |
| Berg et al. (2020) | Data on 52 countries that had reported at least 15 days of at least 100 total confirmed cases and a total of 54 countries that had reported at least 15 days of at least 1 total death from COVID-19. All the analyzed countries had available information on their BCG policies as of **April 1, 2020** | - A linear mixed model of the natural log-transformed number of confirmed cases/ number of deaths controlled for a list of control variables.<br>- Confirmed cases: day 1 is set as the first day of at least 100 confirmed cases.<br>- Confirmed deaths: day 1 is set as the first day of at least 1 confirmed death | Two contrast-coded variables designating BCG policy status:<br>- current vs. [past and none];<br>- combined and past vs. none<br>The effect of BCG policy status on growth rate is reflected by the interactions between day and each BCG policy status contrast | Median age, gross domestic product per capita, population density, population size, net migration rate, and geographical region | - Countries with mandated BCG vaccinations show a significantly slower growth rate of COVID-19 cases and COVID-19 related deaths<br>- Countries that terminated BCG policies before 2000 do not report a growth rate that would be significantly different from countries without BCG policies |
| Dayal and Gupta (2020) | Data on 12 countries that had BCG revaccination policy in the past<br>No exact information which | Comparison of the mean case fatality rates (CFR) between countries with high rate of COVID-19 cases. | Case fatality rates (CFR) | NA | The study finds a significant difference in the COVID-19-related fatality rates (CFR) between with high rate of |
|  | COVID-19 affected countries were selected for the analysis, data collected as of **March 29, 2020** | All countries selected for the analysis followed BCG revaccination policies in the past |  |  | COVID-19 cases and those with BCG revaccination policies |
| Hegarty et al. (2020) | Data on 178 countries dating back 15 days (**March 9-24, 2020**); source accessed on March 24 | The researchers collate all reported global cases and fatalities of COVID-19 and calculate per million incidences and compare them between countries with and without BCG policies in place | Data comparison based on:<br>- reported COVID-19-related cases and fatalities<br>- population numbers<br>- BCG/- non-BCG immunizing countries | NA | The study finds lower levels of COVID-19-related cases and COVID-19 death rates in countries with BCG vaccination as compared to countries with no such a program. |
| Li et al. (2020) | - Data from 167 countries or regions on BCG immunization coverage among 1-year-olds born between 1980 and 2018<br>- Data from 202 countries or regions on COVID-19 daily cumulative cases and cumulative deaths | - T-test to check the difference in the crude growth rate between two groups of countries with at least ten days of data after the cumulative confirmations exceeded 100:<br>Group 1: 39 countries that have no BCG policy for 1980- 1985;<br>Group 2: 52 countries with BCG policy for 1980- 1985.<br>- Univariable linear regression applied on the group 2 dataset that uses the BCG coverage as a factor and either the growth rate or raw case-fatality-rate as response | - COVID-19 growth rate or raw case-fatality-rate as response<br>- BCG coverage as a factor | NA | The study finds no significant effects of BCG coverage on the crude growth rate of COVID-19<br>The conclusion holds both after extension of the 10-day range to 20-day range and lowering the threshold from 100 cases to 50 cases.<br>Also, at the same stage in the epidemic the study does not find any significant difference in COVID-19 severity between countries with or without BCG policies |
| Miller et al. (2020) | - Data on COVID-19 cases and death per country captured on **March 21, 2020**.<br>- Data on BCG policies across | - Analysis of difference of means of COVID-19 cases per million inhabitants (Wilcoxon rank sum test), | The number of deaths per million inhabitants per COVID-19 affected country<br>Only countries with more than | - Countries divided in three categories GNI per capita:<br>1. low income (L), 2. lower middle income, | The study suggests that BCG might show long-lasting protection against COVID-19 |
|  | countries (dates of initiation of BCG vaccination included) | - Linear correlation between the beginning of universal BCG vaccination and the protection against COVID-19 | one million of inhabitants taken into consideration | 3. middle-high- and high-income |  |
| Sala and Miyakawa (2020) | - Data on 136 countries or regions impacted by COVID-19 that includes information on total cases per one million population, life expectancy, and temperature (Model 1).<br>- The data on 91 countries' total deaths per one million population and deaths/cases ratio available (Models 2 and 3)<br>- Data retrieved on **March 28, 2020** | Three sets of <u>linear regressions</u> are applied. DVs: the rate of total cases per one million population (<u>Model 1</u>), the rate of death per one million population (<u>Model 2</u>), and the ratio between these two variables (<u>Model 3</u>).<br>The Order Quantile (ORQ) normalization transformation is used to standardize the continuous variables | - The country's BCG policy with three levels: <u>Group A:</u> countries with BCG policy; <u>Group B:</u> countries with past BCG policy and no BCG policy at the moment; and <u>Group C:</u> countries with no history of BCG policy in place.<br>- Countries divided into three categories: high, middle, or low growth rate of the COVID-19 cases | - Country's life expectancy<br>- The average temperature in February and March 2020 | The study finds that the number of total cases and deaths per one million population are significantly associated with the country's BCG policy |
| Shet et al. (2020) | Data on top 50 countries reporting highest COVID-19 case events with at least 100 reported cases and adjusted for time since the event (days since 100th case reported as of **29 March 2020**) | - Simple log-linear regression model to assess the association of BCG use and COVID-19-related mortality per 1 million population<br>- The Shapiro-Wilk normality test to confirm that the log-transformed COVID-19-specific mortality is normally distributed | - COVID-19-attributable mortality data per 1 million population for each country,<br>- BCG policy in place as exposure | - GDP per capita,<br>- The percentage of population 65 years and above,<br>- The relative position of each country on the epidemic timeline | The study finds that COVID-19-related mortality among countries with BCG policy in place is lower than in countries that do not have BCG policy |

The papers presented above analyze the relation between BCG vaccinations and either COVID-19-related case events and fatalities (e.g., Li et al., 2020, Hegarty et al., 2020, Sala and Miyakawa, 2020, Shet et al., 2020) or only COVID-19-related fatality rates (Dayal and Gupta, 2020). The majority of the studies use publicly available data repositories such as Worldometer to source COVID-19-related data. In the same time most studies source country wise BCG-related data using outlets such as BCG World Atlas (http://www.bcgatlas.org/). Other sources, such as reports and datasets published by World Health Organization, World Bank and United Nations are also used to retrieve economic and demographic information for countries under investigation.

Below we present most important highlights and findings related to the analyzed literature. Shet et al. (2020) apply simple log-linear regression model to investigate COVID-related mortality and BCG vaccinations. The researchers use the following covariates in their model: GDP per capita, the percentage of population 65 years and above and the relative position of each country on the epidemic timeline. The findings show that countries with BCS vaccinations show lower levels of COVID-19-related mortality. In turn, Berg et al. (2020), after analyzing data on first 30-day COVID-19 country-wise outbreaks, argue that BCG immunizations are associated with flattened growth curves for confirmed cases of COVID-19 infections and deaths. Similarly, Hegarty et al. (2020) argue that countries that vaccinate their populations against BCG, experience less COVID-19-related cases and lower death rates caused by COVID-19. Miller et al. (2020) propose that differences between countries in COVID-19 impact could be partially explained by different levels of BCG policy implementations.

In summary, the extant studies find that there is sufficient empirical evidence to show countries that have BCG coverage in NIPs have significantly lower case and death rates based on available data till April 01, 2020. Further, none of these studies take into account heterogeneity in testing rates across countries.

### Methodology

We used Worldometer to collect national COVID-19 attributable data as of April 10, 2020 which includes ‘cases per million’ and ‘deaths per million’ attributes for the top 115 countries reporting highest case events. We used the same source to collect information on COVID-19 testing data at country level. We used the data to derive other variables such as ‘cases per test’ and ‘deaths per case’, which represented country-specific data on the number of COVID-19 positive cases per conducted tests and the probability of death for an individual who tested positive for COVID-19. We included data on countries’ economic status by collecting country-related Human Development Index (HDI) score and Corruption Perception Index (CPI) score. We derived country population and economic information for 2018 (GDP per capita and income status for respective countries) from the World Bank population data. We controlled for the relative population density of a country by including ‘population density per km^2^ along with the ‘percentage of urban population within total population’ score. Similar to prior studies (Shet et al., 2020) we calculated two parameters to align the countries on a more compatible time curve to mitigate bias centered due to differential epidemic time curves experienced by different countries. The two variables are as follows:

1. The number of days since a country has reached 100 cases and
2. The number of days since a country has reached 1 case per million.

We included data on BCG vaccine inclusion in national immunization schedules from the BCG World Atlas. We categorized the countries in three groups:

- Group 0: 5 countries which have never included BCG vaccine in their schedule;
- Group 1: 21 countries which had BCG vaccine included in the past but do not have it currently and
- Group 2: 72 countries which currently include BCG vaccine in their national immunization schedules

We also created a BCG Index variable which represents the total number of years since 1950 during which a country mandated BCG vaccine inclusion in national immunization schedules. For example, India mandated BCG vaccination in NIP in 1948 and continues to have it today, so BCG index for India is 1. Spain introduced BCG coverage in 1965 and discontinued it in 1981, so BCG index for Spain is 16/70.

To evaluate the effect of BCG vaccine on either the total amount of infection (the total number of cases per million) or the mortality rate (deaths per million), we created multiple regression models. We performed the analysis for the following four dependent variables (DVs): ‘the total number of cases per million’, ‘total number of deaths per million’, ‘the total number of deaths per case’ and ‘the total number of cases per test’. To evaluate the effect of BCG vaccine on the discussed dependent variables, we ran separate models using continuous BCG Index variable or categorical BCG variables: Group 0, Group1 and Group 2 as described above.

We focused on the analysis of data from 98 countries. We controlled for the effects of the following variables: GDP per capita, HDI score, population density per km2, percentage population of above 65 years of age, government corruption perception index, total percentage of urban population, total number of days since 100 cases and total number of days since 1 case per million as of 10^th^ April 2020. We excluded countries that lacked the information about their BCG inclusion policies or whose population was less than 1 million. For the countries with a BCG vaccination in place which lacked the information on the start date and end date of the BCG vaccination policy, we approximated the number of years of coverage to be 35 years.

We used R and Python to perform the analysis and we present our results below.

## Results

We present the comparison of the mortality (number of deaths per million) among groups of countries with differing levels of inclusion of BCG vaccine in Figure 6. Group 2 countries reported a significantly lower count of deaths per million than countries belonging to Group 0 and Group 1. As pictured in Figure 6, we observed similar trends for the total number of cases per 1 million population, number of deaths per case and the number of cases per test conducted.

**Figure 2:**
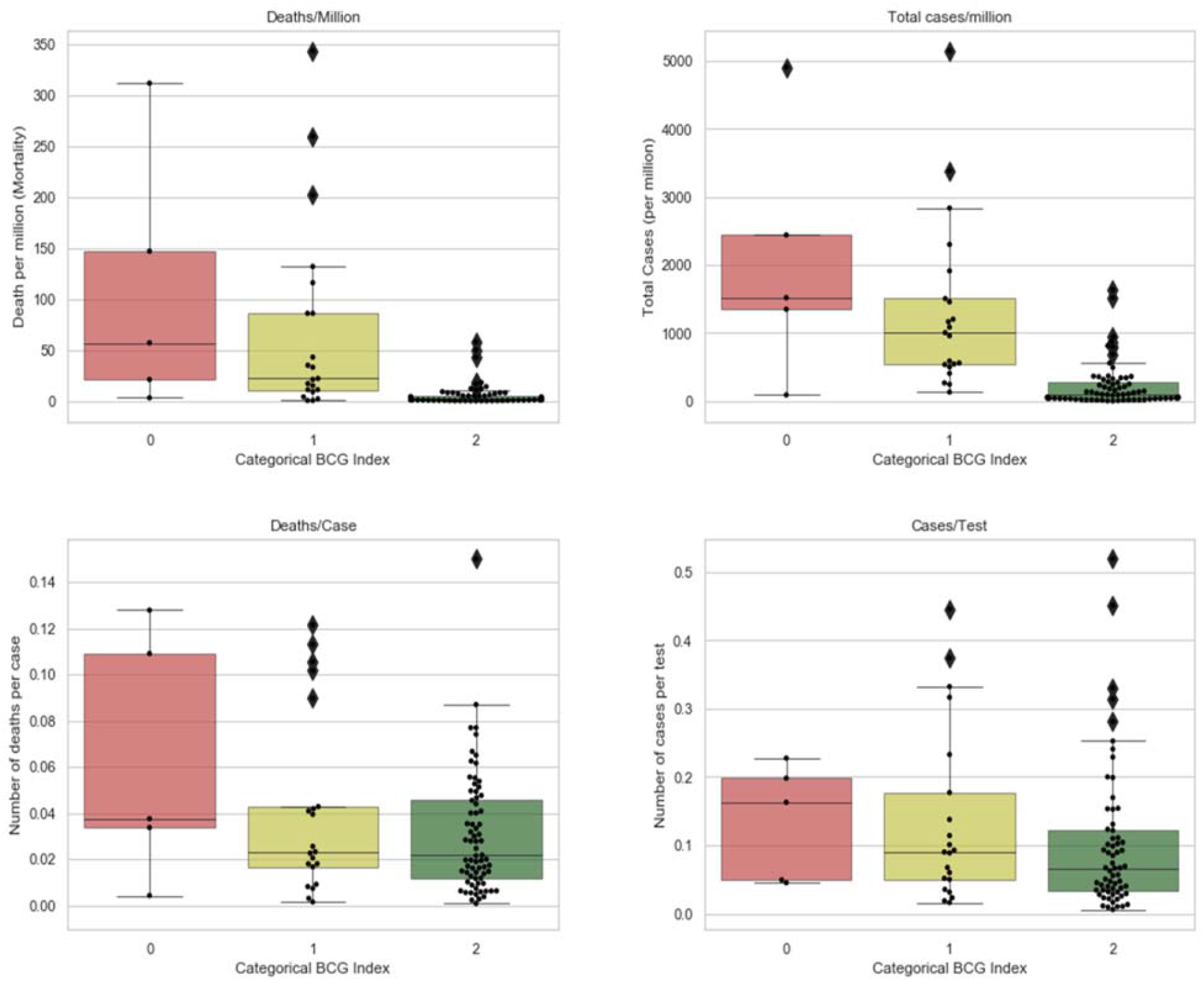
**Box plots for cases per million, deaths per million, number of deaths per cases and number of cases per test as per BCG inclusion level for countries**

We collected descriptive statistics of infection rates, mortality rates and the number of tests per million to capture the heterogeneity present in data (Table 2). We observed that as compared to the other variables the standard deviation of the attribute ‘tests per million’ was significantly higher and accounted for 13473 tests. It demonstrated a significant heterogeneity related to the amount of tests conducted by countries.

**Table 2:** Descriptive statistics of case incidence, mortality and testing data.

|  | Total Cases per Million | Deaths per Million | Tests per Million | Deaths per Case | Cases per Test |
| --- | --- | --- | --- | --- | --- |
| Min | 1 | 0.03 | 13 | 0.0008584 | 0.00567 |
| 1st Quantile | 43.25 | 1 | 586.5 | 0.0119881 | 0.038 |
| Median | 186 | 3 | 2884.5 | 0.0239569 | 8883 |
| Mean | 547.94 | 24.25 | 7501.5 | 0.0344656 | 0.16953 |
| 3rd Quantile | 558 | 12 | 10496.8 | 0.0463377 | 0.1928 |
| Max | 5149 | 344 | 100002 | 0.15 | 1 |
| Std. Dev. | 922.06 | 60.3 | 13473.2 | 0.0314 | 0.24 |

In the regression models we tested the effect of BCG categorical or BCG Index on our dependent variables, controlling for other covariates such as GDP per capita, HDI score, CPI score, population density, percentage of above 65 population, urban population percentage, number of days since 100 cases and number of days since 1 case per million. We included another control variable – ‘test per million’ for three dependent variables. We excluded ‘test per million’ for the ‘cases per test’, as the two variables are highly correlated.

To evaluate the effect of categorical BCG values on COVID-19 cases per million population, we observed that countries belonging to Group 2 had on average 916 less cases per million than countries represented by Group 0. Group 1 countries did not have a statistically significant difference in case incidences per million as compared to the countries represented by Group 0.

Results for ‘deaths per million’ were similar, as countries with current BCG on an average have 93 deaths less per million as compared to countries which never had BCG as part of their NIP. For the dependent variable ‘deaths per positive case’ Group 2 countries had 4 percent less deaths per every positive case than countries belonging to Group 0. No such difference was observed between Group 1 and Group 0.

Adding the testing component, we then used the dependent variable ‘cases per test’, and observed that there was no effect of BCG inclusion in a countries’ national immunization program on the number of COVID-19 cases it reported conditional on tests conducted. Our findings are presented in Table 3.

**Table 3:** **Linear regression model of COVID-19-attributable case incidence/ mortality variables and BCG use in national immunization schedules, adjusting for relevant covariates. Exponentiated estimates for the parameters are provided**.

|  | Cases Per million | Deaths per million | Deaths per case | Cases per test conducted |
| --- | --- | --- | --- | --- |
| BCG currently not used in NIP | -322.6 | -40.47 | -0.02261 | 0.008961 |
| BCG used in NIP | -916.8*** | -93.45*** | -0.04259** | 0.01742 |
| GDP per capita (PPP) | 0.01642*** | 0.0007629 | 5.396e-08 | 1.954e-06 |
| Above 65 population percentage | 40.64** | 3.006* | 0.001112 | 4.396e-03 |
| Number of days since 100<sup>th</sup> case | 8.68 | 1.912* | 0.001366** | 2.132e-03 |

When we replaced the categorical variables capturing BCG inclusion in countries’ NIP with the continuous variable BCG Index, we observed that the BCG Index has a strong significance on the number of COVID-19 cases per million. With each additional year of BCG vaccine inclusion in a country’s NIP, we observed a reduction of 13 COVID-19-related cases per million population. Similarly for mortality, each additional year of BCG inclusion resulted in one less death per million. For ‘deaths per case’ in population, BCG Index was found to be significant, with each additional year of BCG inclusion being responsible for 0.003487 less deaths per case. For ‘cases per test’ variable, the effect of BCG Index was found to be insignificant. Our findings are provided in Table 4.

**Table 4:**
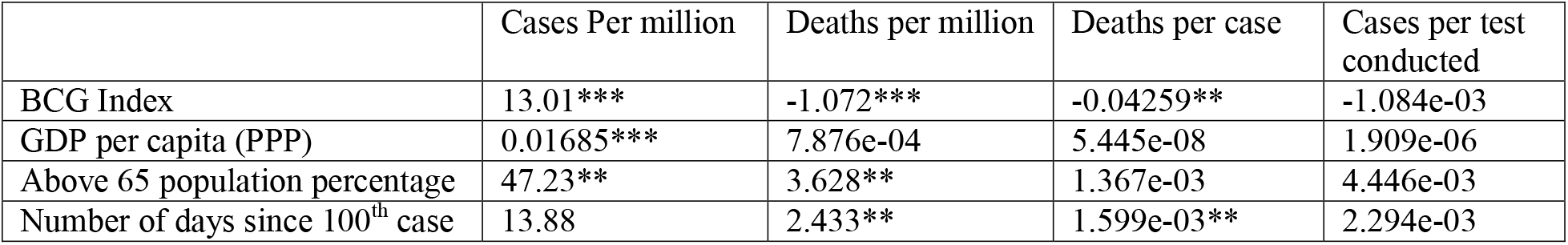
**Linear regression model of COVID-19-attributable case incidence/ mortality variables and BCG use in national immunization schedules, adjusting for relevant covariates. Exponentiated estimates for the parameters are provided**.

Log-linear regression models have also been used by researchers in the past (Shet et al., 2020). We created multiple log linear models where we applied logarithmic transformation on the following variables: ‘GDP per capita’, ‘tests per million’ and ‘cases per million’. The log-linear regression results are consistent with results reported in Tables 3 and 4. Categorical BCG Group 2 and BCG Index were found to be statistically significant factors for ‘cases per million’, ‘deaths per million’ and ‘deaths per case’. However, both the categorical and continuous BCG inclusion variables were found to be insignificant for the ‘cases per test’ variable in the log linear models as well.

## Discussion

Recently published studies have conducted multiple analyses and found that the inclusion of BCG vaccines in a country’s NIP to have a significant effect on COVID-19 infection cases and deaths. Although our study shows similar findings, when we do not take into account testing, we find that the coefficient estimates of the BCG-related variables have reduced in value based on the analysis on the latest data from April 10, 2020. We observed high heterogeneity in the amount of testing conducted by various countries which adds an interesting dimension to consider while looking at the relationship between BCG vaccine inclusion in a country’s NIP and the rate of COVID-19 infection and mortality.

Compared to the amount of testing conducted by countries, case incidence or mortality variables demonstrated significantly lower level of heterogeneity. As the detection of infection is greatly dependent upon the amount of testing being done, we believe there may be endogenous factors affecting the relationship between the inclusion of BCG vaccine in a country’s NIP and COVID-19 infection and mortality rates. In the current scenario, testing is the only mechanism through which COVID-19 cases are identified. As such it stands to reason that if a country does not test extensively, it will also report significantly lower number of COVID-19 cases and deaths. Therefore, our dependent ‘Cases per test’ captures the incidence of disease among those for which measurement has been taken. The robust result of non-significance of BCG inclusion in NIP raises questions on the validity of the assertion that BCG vaccine effects the respective spread of COVID-19 in a population. Hence, we believe the since when testing for COVID measure is included in the analysis, the impact of BCG policy on infection rates vanishes, there is no statistical evidence to support the assertion that BCG vaccination policy lowers infection rate at the country level.

### Conclusion and Future Work

The landscape of COVID-19 attributable deaths and cases throughout the world is constantly evolving. Given this constant change, we have to be careful before making assertions on how BCG vaccinations influence the emergence of COVID-19-related cases and deaths. We cannot exclude endogenous factors such as testing, that affect the relationship between the BCG vaccine and number of COVID-19 attributable cases and mortality. We argue that it may be beneficial to create a longitudinal study that can be used to get more insights of the relationship between the BCG vaccine and COVID-19. We are currently in the process of creating a panel dataset to analyze the effects of the BCG vaccine inclusion on COVID-19 attributable cases and mortality change over time.

## Data Availability

The data is taken from public sources, some of whom are: https://www.worldometers.info/coronavirus/ accessed on April 10 at 8.00 pm (EST).
https://en.wikipedia.org/wiki/List_of_countries_by_Human_Development_Index
Government transparency score can be considered as a proxy for reliability of reported COVID 19 data at the country level (Hollyer et al 2014).

## Footnotes

1 https://www.worldometers.info/coronavirus/ assessed on 04/18 at 8.15 pm.

2 https://www.worldometers.info/coronavirus/ accessed on April 10 at 8.00 pm (EST).

3 https://en.wikipedia.org/wiki/List_of_countries_by_Human_Development_Index

4 Government transparency score can be considered as a proxy for reliability of reported COVID 19 data at the country level (Hollyer et al 2014).

